# Serum soluble urokinase plasminogen activator receptor as a biomarker for distinguishing Kawasaki disease from infections in febrile children

**DOI:** 10.1101/2023.01.26.23285054

**Authors:** Ahmed R. Alsuwaidi, Junu A. George, Hassib Narchi

## Abstract

**Background:** The soluble form of the urokinase plasminogen activator receptor (SuPAR) is a potential biomarker in various inflammatory, infectious, and autoimmune conditions.

**Objectives:** In this stusy, we aimed to evaluate its diagnostic utility in febrile children to distinguish between Kawasaki disease (KD) and infections, and to investigate any association with the development of coronary artery aneurysms (CAA) KD.

**Methods:** In this retrospective observational cohort study we enrolled 17 children with fever lasting more than 5 days and without suggestive diagnostic signs on admission to hospital. Serum SuPAR concentrations were measured on admission and compared between children with confirmed KD and those with infections, as well as between children with KD who did or did not develop CAA.

**Results:** KD was later confirmed in seven children (median age 25 months), and febrile infections in 10. There was no significant difference in suPAR concentrations between both groups: 5.35 ± 2.76 ng/mL in KD, and 5.57 ± 1.69 ng/mL in febrile infections (*p*=0.84). The best cut-off value for suPAR, ≥ 7.74 ng/mL, was the best to correctly classify 64.7% of the cases, with a sensitivity of 28.6% and specificity of 90%. However, it had a low diagnostic performance (Youden index 18.6%, area under the curve curve 60%), and therefore failed to differentiate between KD and infections. In the seven children with KD, only one child developed CAA (SuPAR 4.69 ng/mL) while six other did not (SuPAR 5.47 ± 1.04 ng/mL) but the statistical significance could not be computed.

**Conclusion:** In febrile children, serum suPAR concentrations failed to distinguish between KD and infections, and were not associated with the development of CAA in KD. Therefore, SuPAR is not a useful biomarker in the diagnosis or prognosis of KD.

## INTRODUCTION

Urokinase plasminogen activator receptor (uPAR) binds to the cell membrane of immunologically active cells, such as monocytes, activated T-lymphocytes, macrophages, endothelial, and other cells. It separates from the cell during inflammation, generating its soluble form, suPAR, which can be measured in blood and other body fluids. It facilitates the passage of inflammatory cells from the bloodstream into tissues and plays a role in the plasminogen-activating pathway, inflammation, and modulation of cell adhesion, migration, and proliferation. As the urokinase receptor system regulates the confluence of inflammatory, immune, coagulation, and fibrinolytic responses, suPAR has been used in the diagnosis and prognosis of multiple inflammatory, infectious, autoimmune, neoplastic conditions, as well as coronary heart disease and heart failure.^1-10^ Regardless of the underlying pathology, serum levels are raised in acutely ill patients, and also have a prognostic value as they predict high mortality.^6^ As the biochemical and molecular mechanisms underpinning these properties remain unclear, suPAR is currently considered a non-specific biomarker for disease presence, progression, and severity.^6^

Kawasaki disease (KD) is an acute febrile vasculitis of unknown etiology that affects mainly infants and young children, some of whom develop coronary artery aneurysms (CAA) if not treated early with intravenous immunoglobulin (IVIG).^11^ Activation of the innate immune system induces vasculitis and triggers the adaptive immune system to produce cytokines, chemokines, proteases, and reactive oxygen species.^12^ Further recruitment of immune cells into the arterial wall ensues, where inflammation and oxidative stress interact and amplify each other, resulting in endothelial dysfunction.^13^ KD needs to be differentiated from more common febrile illnesses, as early diagnosis is essential in preventing CAA. but, without pathognomonic diagnostic tests, its diagnosis remains essentially clinical.^11^ Thus, laboratory investigations that helps in its diagnosis remain highly sought after.

We hypothesized that the complex inflammatory processes that underlie vasculitis in KD may result in overexpression of suPAR, and make it a potentially useful biomarker. To the best of our knowledge, this hypothesis has not been explored before. To test this hypothesis, we compared suPAR concentrations between children with KD and those with other febrile infections. Additionally, we investigated whether suPAR concentrations were associated with the development of coronary artery aneurysms (CAA) in children with KD..

## METHODS

This retrospective observational cohort study is a continuation of an earlier project.^14^ It was performed on preserved residual sera from 17 children under six years of age who had fever for ≥ five days and without suggestive diagnostic signs on admission.

Plasma samples were collected within 24 hours of admission and were immediately transported to the laboratory for storage at −80°C until further analysis. Plasma suPAR levels were measured using commercial Enzyme-Linked Immunosorbent Assay (ELISA) kits (suPARnostic™ assay, Virogates, Copenhagen, Denmark). Plasma (25 μL) was mixed with 225 μL of horseradish peroxidase (HRP)-labelled detection antibody. A total of 100 μL of this mix was then transferred to duplicate wells of a microwell plate precoated with capture anti-suPAR antibody. The plates were washed after one hour of incubation at room temperature. To each well, 100 μL of substrate was added and incubated for 20 minutes in the dark before color development was stopped by the addition of sulphuric acid. The optical absorbance was measured at 450 nm using a microplate reader. Plasma suPAR concentration (expressed as ng/mL) was determined by interpolation of the calibration curve prepared from the provided suPAR standard. The lower limit of detection of the assay was 0.1 ng/mL as determined by the manufacturer.

Serum suPAR concentrations, normally distributed continuous variables, were reported as mean ± standard deviation and were compared with the unpaired Student t-test. Sensitivity, specificity, Youden index (sensitivity + specificity -1), positive and negative likelihood ratios, as well as the receiver operating characteristic (ROC) of SuPAR concentrations to predict KD were calculated. The cut-off concentration to differentiate between the two groups was that with a Youden index of 50%, with values below demonstrating failure to differentiate KD from febrile infections. All analyses were performed using Stata version 16 (Stata Corp, Texas) and a two-tailed *p*-value <0.05 defined statistical significance.

## RESULTS

There were 17 children (median age 25 months; interquartile range 18, 53 months) including seven boys. KD was diagnosed in seven patients (of whom only one developed CAA) and ten had febrile infections (one bacterial pneumonia, one scarlet fever, three viral respiratory infections, and five viral gastroenteritis). Intravenous immunoglobulins were administered to all the children with KD. Their full details were published earlier.14

There was no significant difference in suPAR concentrations between children with KD (5.35 ± 2.76 ng/mL) and febrile infections (5.57 ± 1.69 ng/mL) (p=0.84) (Fig. 1 Panel A). Additionally, SuPAR concentrations were not significantly different between bacterial and viral infections (4.75 ± 1.07 ng/mL vs 5.78 ± 1.81 ng/mL) (p=0.47), between sexes (p=0.61), age groups (p=0.47) or duration of fever before admission (p=0.61). Similarly, they showed no correlation with the highest recorded temperature (p=0.28) or serum C-reactive protein level (p=0.45).

**Figure 1.**
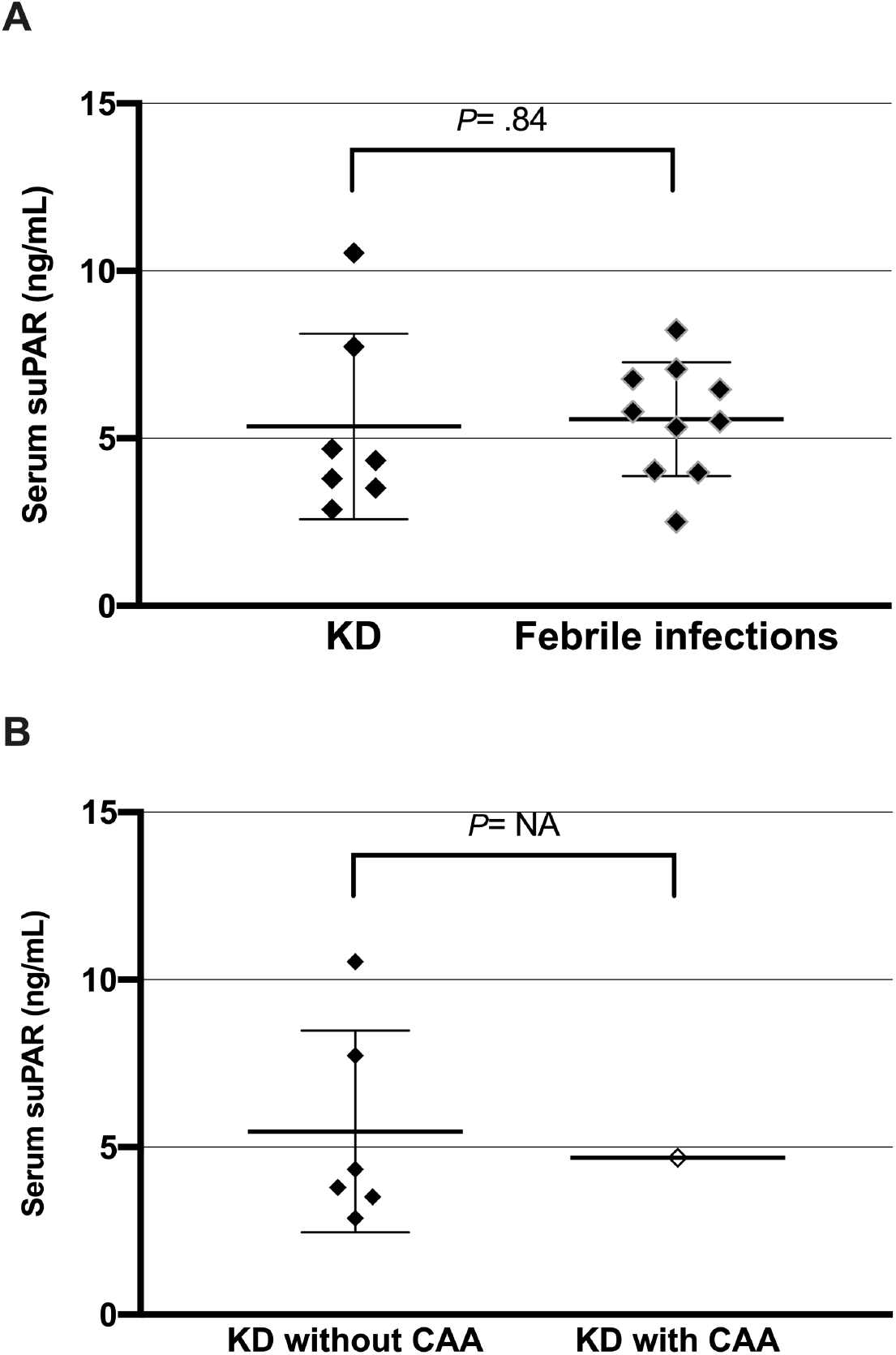
Soluble Urokinase Plasminogen Activator Receptor (SuPAR) concentrations in 7 children with Kawasaki disease (KD) and 10 with febrile infections. *Panel A:* KD versus febrile infections. *Panel B:* KD with and without coronary artery aneurysms (CAA). Horizontal lines represent mean values ± standard deviation. *p* values calculated by the 2-sided unpaired Student’s t-test.

The performance of suPAR concentrations in predicting KD included calculation sensitivity, specificity, Youden index, positive and negative likelihood ratios (Table 1). The optimal cut-off point for suPAR concentrations, with the highest Youden index of 18.6%, was ≥ 7.74 ng/mL, which correctly classified 64.7% of cases., with a sensitivity of 28.6% and specificity of 90%. However, Youden index was < 50%, and the surface area under the curve was 60%. (Figure 2).

**Table 1.** Characteristics of serum suPAR concentrations in Kawasaki disease and febrile infections in 17 children with fever.

| suPAR concentration<br>cutpoint (ng/mL) | Sensitivity | Specificity | Correctly<br>classified | LR+ | LR- | Youden<br>index |
| --- | --- | --- | --- | --- | --- | --- |
| ≥ 3.8 | 71.43% | 10.00% | 35.29% | 0.7937 | 2.8571 | -18.57% |
| ≥ 3.99 | 57.14% | 10.00% | 29.41% | 0.6349 | 4.2857 | -32.86% |
| ≥ 4.04 | 57.14% | 20.00% | 35.29% | 0.7143 | 2.1429 | -22.86% |
| ≥ 4.34 | 57.14% | 30.00% | 41.18% | 0.8163 | 1.4286 | -12.86% |
| ≥ 4.69 | 42.86% | 30.00% | 35.29% | 0.6122 | 1.9048 | -27.14% |
| ≥ 5.34 | 28.57% | 30.00% | 29.41% | 0.4082 | 2.381 | -41.43% |
| ≥ 5.51 | 28.57% | 40.00% | 35.29% | 0.4762 | 1.7857 | -31.43% |
| ≥ 5.8 | 28.57% | 50.00% | 41.18% | 0.5714 | 1.4286 | -21.43% |
| ≥ 6.46 | 28.57% | 60.00% | 47.06% | 0.7143 | 1.1905 | -11.43% |
| ≥ 6.78 | 28.57% | 70.00% | 52.94% | 0.9524 | 1.0204 | -1.43% |
| ≥ 7.07 | 28.57% | 80.00% | 58.82% | 1.4286 | 0.8929 | 8.57% |
| **≥ 7.74** | **28.57%** | **90.00%** | **64.71%** | **2.8571** | **0.7937** | **18.57%** |
| ≥ 8.24 | 14.29% | 90.00% | 58.82% | 1.4286 | 0.9524 | 4.29% |

LR+ and LR- : positive and negative likelihood ratio; Youden index= sensitivity + specificity -1
The cut-off point of serum suPAR concentrations $\geq 7.74$ ng/mL (highlighted in bold), had the highest Youden index (18.6%) and was the best to correctly classify 64.7% of the cases with a sensitivity of 28.6% and specificity of 90%.

**Figure 2.**
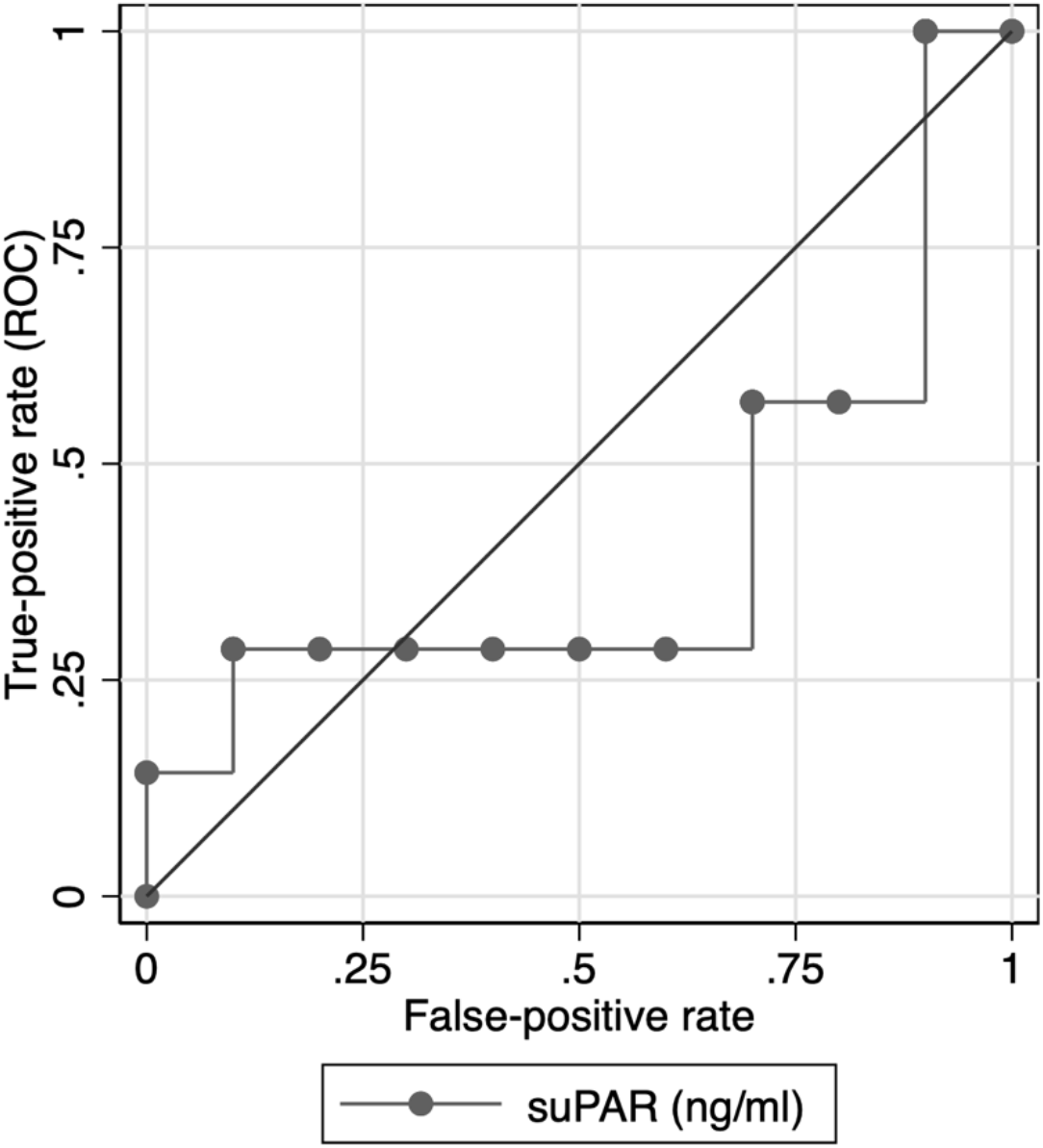
Receiver Operating Characteristics (ROC) curve of soluble urokinase plasminogen activator receptor (SuPAR) concentrations to differentiate between Kawasaki disease and febrile infections.

One child with KD developed CAA and had a suPAR concentration of 4.69 ng/mL, compared to 5.47 ±1.04 ng/mL in the other six children (Figure 1, panel B). The *p-*value could not be computed.

## DISCUSSION

To the best of our knowledge, this is the first study to investigate suPAR concentration in children with KD. Although serum levels were elevated in both KD and febrile infections, the difference was not statistically significant. Furthermore, the performance of SuPAR to diagnose KD was poor, as the optimal cut-off value ≥ 7.74 ng/mL with the highest Youden index of 18.6%, was well below 50%, and the surface area under the curve was only 60%. It is not surprising that suPAR levels would be similar in children with KD and febrile infections, as both conditions can cause systemic inflammation.^3,5,15^

In KD patients, there was also no difference in suPAR levels in the presence or absence of CAA. Reasons for this include the small sample size of the cohort with only one child with KD developing CAA. Additionally, suPAR levels increase in KD where the vasculitis may extend to the coronary arteries. Thus, it is not surprising that suPAR levels would be similarly elevated in both KD and its CAA complication.

In addition to the limitations mentioned erlier, others include its retrospective nature, and the measurement of a single suPAR concentration only upon admission to hospital. To confirm the findings of this stusy, larger, prospective, multicenter studies with larger sample sizes and serial suPAR measurements are needed. Exploring the potential of combining SuPAR measurement with other biomarkers or clinical factors to improve diagnostic accuracy in KD are also indicated.

## CONCLUSION

This study found that serum suPAR concentrations on admission to the hospital were not able to differentiate between KD and infections in febrile children. Additionally, they were also unable to distinguish between KD patients who developed CAA and those who did not.

## Data Availability

All data produced in the present work are contained in the manuscript

## Acknowledgments

We are grateful to the participating children, their families, and the staff who cared for the them.

## Contributors

HN and ARA conceptualized and designed the study, and JG performed the laboratory analyses. HN analyzed the data and drafted the initial manuscript. All authors critically reviewed and revised the manuscript for important intellectual content, and approved the final manuscript as submitted, and agreed to be accountable for all aspects of the work.

## Ethics approval

The study was performed in accordance with the Declaration of Helsinki of the World Medical Association and was approved by the United Arab Emirates University Human Research Ethics Committee under registry number ERH-2021-7392

## Patient consent for publication

The parents of all the participating children signed an informed consent form for this study.

